# Classification of type 1 diabetes and type 2 diabetes based on administrative registry data: A nationwide study from Norway

**DOI:** 10.64898/2025.12.05.25341685

**Authors:** Paz Lopez-Doriga Ruiz, Lars C. Stene, Karianne Fjeld Løvaas, Grethe Åstrøm Ueland, Tone Vonheim Madsen, Kåre I Birkeland, Hanne Løvdal Gulseth, Robert Wiik, Kjersti Nøkleby, Inger Johanne Bakken

## Abstract

**Background:** Type 1 diabetes (T1D) and type 2 diabetes (T2D) differ in health care needs, and precise prevalence estimates by diabetes type are important for health care planning.

**Data and Methods:** Using Norwegian health registry data on diagnoses from primary and specialist care and prescribed drugs, we developed a classification method to define T1D and T2D. The Norwegian Diabetes Register for Adults (NDR-A) served as reference standard.

**Results:** First, for individuals in the NDR-A, we established a health registry data classification method with three key steps: (A) classifying T1D based on the use of insulin and diagnoses from specialist healthcare, (B) classifying T2D based on diagnoses from both specialist and primary healthcare, and (C) identifying individuals as having T1D if they were diagnosed in primary healthcare and used insulin, but never had a T2D diagnosis. Overall correspondence between NDR-A diagnosis and the classification method was 94.7% for T1D and 97.9% for T2D. For T1D, the correspondence was 97% or higher in age-groups 18-39 and 40-59, 87.6% for those aged 60-79 and 69.4% for individuals older than 80 years. For T2D, the correspondence was 97% or higher for age-groups 40 and above, and 92.1% for those aged 18-39. Applying the classification method to total population data showed that, in 2022, among 4,315,563 individuals aged 18 years and older living in Norway, 31,264 were identified as having T1D (0.72%) and 239,120 were identified as havingT2D (5.5%).

**Conclusion:** While classifying diabetes types can be challenging, our registry-based classification corresponded well with the NDR-A.

## Introduction

Diabetes is primarily classified into two major types: type 1 diabetes (T1D) and type 2 diabetes (T2D). T2D comprises more than 85% of all cases, while T1D accounts for approximately 5-10%, with other forms remaining relatively rare [1]. The two main diabetes types differ in pathophysiology, risk factors, prognosis, management, and health care needs [2]. Notably, T1D is much more common than T2D in children, whereas the incidence and prevalence of T2D increases markedly with age, at least up to 80 years. Accurate estimates of diabetes prevalences are important for effective health system planning. Robust public health surveillance can be achieved through prospectively collected individual-level data [3]. In registry-based research, diabetes type is usually defined using diagnosis-related codes and/or information on the use of blood-glucose lowering drugs [4]. However, the accuracy of these methods is often unknown [3].

Overlapping treatment strategies can complicate accurate classification of diabetes type. T1D requires treatment with insulin, yet insulin is also prescribed to a subset of individuals with T2D. Similarly, not all patients with T2D are treated with glucose-lowering drugs [5], further complicating classification based on prescription data alone [5-7]. Age at diagnosis has been employed as a criterion in some studies to distinguish between T1D and T2D [8]. Research on T1D has predominately been focused on paediatric populations, whereas studies on T2D have primarily been limited to adults. However, this approach could lead to bias, as recent findings indicate that approximately half of all patients with T1D are diagnosed as adults [9]. Furthermore, in Norway, 10% of all T2D cases are diagnosed before age 40 [10].

According to the Norwegian national diabetes guidelines, individuals with T1D are offered multidisciplinary follow-up in the specialist health service, and usually have regular follow-up visits in public hospital outpatient clinics. In contrast, the majority of individuals with T2D are diagnosed and managed within primary care settings [11]. The overlapping treatment strategies and diverging care pathways challenge accurate classification of diabetes type when relying on a single registry that includes only information on prescriptions, primary care, or secondary care.

The Norwegian nationwide administrative health registries have a high level of completeness [12, 13], and are extensively used for research [14]. In previous work, we have estimated incidence and prevalence of diabetes by using nationwide registry-linkages covering prescribed drugs and diagnostic codes from both primary and specialist healthcare [15, 16].

In the current study, our primary objective was to utilize data from a medical quality registry for diabetes in adults as a reference point for developing classification strategies. Although this registry contains clinically gathered information and can presumably serve as a gold standard for determining diabetes type, it does not have complete national coverage. We also aimed to apply our classification to estimate the prevalence of diagnosed T1D and T2D diabetes among adults living in Norway in 2022, using nation-wide health registry data.

## Material and methods

All analyses were performed using R Statistical Software (v4.1.2; R Core Team 2021) [23].

### Setting and study population

Data from Norwegian health and population registries can be linked by the unique personal identification number. We used data from the Norwegian Population Registry to identify all individuals 18 years and older residing in Norway during 2022 (N= 4,315,563). The design and data sources are outlined in Figure 1.

**Figure 1.**
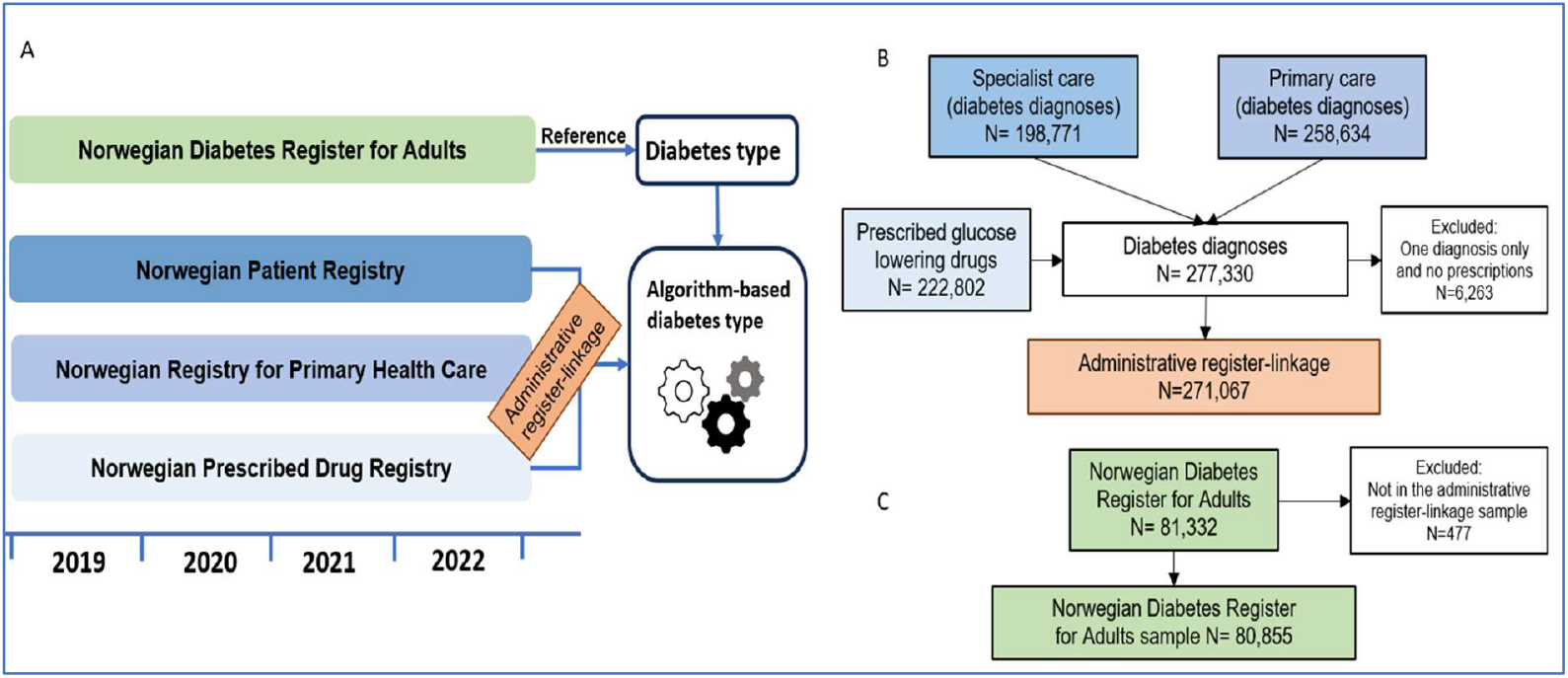
Inclusion flow charts based on data from 2019-2022. A) Study data sources with timeline. B) Administrative registry-linkage data. C) Norwegian Diabetes Register for Adults (NDR-A). NPR: Norwegian Patient Registry; NRPHC: Norwegian Registry for Primary Health Care; NorPD: Norwegian Prescribed Drug Registry. Included data NDR-A: Diabetes type; NPR: ICD-10 codes “E10 Type 1 diabetes mellitus” and “E11 Type 2 diabetes mellitus”; NRPHC: ICPC-2 codes “T89 Diabetes type 1” and “T90 Diabetes type 2”; NorPD: ATC codes “A10A Insulins and analogues” and “A10B Blood glucose lowering drugs, excluding insulins”.

### Data sources

The Norwegian Diabetes Registry for Adults (NDR-A) was established in 2006 as a national medical quality registry [17, 18]. The registry gathers data through forms completed for individuals with diabetes 18 years and older, during two types of patient visits: 1) with general practitioners and 2) with specialists at diabetes outpatient clinics. The aims of the registry are to enhance the quality of diabetes care by providing feedback to physicians and clinics regarding treatment standards, as well as to encourage the use of its data in research [18]. Since 2020 written consent is not required to include individuals in the NDR-A registry, and the number of individuals registered has increased as a result [18]. In 2022, the estimated coverage for T1D and T2D was 88% and 32%, respectively [19, 20]. We used the diabetes classification in the NDR-A as reference (“gold standard”).

General practitioners are reimbursed for each patient encounter through the Norwegian Health Economics Administration (Helfo). This system contains data dating back to 2006, with diagnoses coded according to the International Classification of Primary Care version 2 (ICPC-2). From July 2016 onwards, the data have been reported to the Norwegian Register for Primary Health Care (NRPHC) [13]. This registry was established to facilitate administration, management, financing, and quality assurance of the primary health and care services, as well as to support research. We previously used data directly from the Helfo system [15] but are now using data from NRPHC, as this is the current recommended source for primary health care registry research.

The Norwegian Patient Registry (NPR) includes individual-level data from 2008 onwards [13]. The registry contains data on hospitalizations and outpatient visits, with diagnostic information coded according to the International Classification of Diseases version 10 (ICD-10).

The Norwegian Prescribed Drug Registry (NorPD) was established in 2004 and consists of individual-level information on all dispensed drugs at Norwegian pharmacies, categorized according to the Anatomical Therapeutic Chemical (ATC) classification [21].

The Norwegian Population Registry holds demographic information on all residents of Norway, such as sex and dates of birth, death, immigration, and emigration [22].

### Variables

From the NDR-A, we collected information from 2019 to 2022 on the diabetes type for all individuals registered as alive and residing in Norway in 2022 in the Norwegian Population Registry. We included individuals diagnosed with T1D and T2D; other diabetes types were excluded. Note that latent autoimmune diabetes in the adult (LADA) is classified as T1D in the NDR-A.

From the NPR, we extracted information on all hospitalizations and outpatient visits where the ICD-10 codes E10 (“*Type 1 diabetes mellitus*”) or E11 (“*Type 2 diabetes mellitus*”) were registered from 2019 to 2022. Similarly, from the NRPHC, we extracted information on all primary care contacts with ICPC-2 codes T89 (“*Diabetes type 1*”) or T90 (“*Diabetes type 2*”) were registered during the same period.

From the NorPD, we extracted data on all dispensed drugs with ATC codes A10A (insulins and analogues) and A10B (blood glucose lowering drugs excluding insulins) during 2019-2022. We grouped ATC codes for blood glucose lowering drugs as follows (note that group 1b is a subset of group 1a):

1. Insulins and analogues:
  a. All A10A (insulins and analogues)
  b. A10AB (fast acting insulins and analogues)
2. Blood glucose lowering drugs, excluding insulins: A10B

We constructed count variables indicating the number of registrations with each diagnostic code (E10, E11, T89, T90) or within each ATC group (A10A, A10AB, A10B).

### Classification of diabetes type

The NDR-A diagnosis was used as the reference for classification of diabetes, while the administrative registry-linkage was considered to have complete population coverage for all individuals with diabetes.

We aimed at distinguishing between T1D and T2D in all age-groups 18 years or older, keeping the procedure as simple as possible. We defined the criteria for inclusion as having at least two entries of a diagnosis code for diabetes or at least one entry of a diagnosis code combined with a dispensed prescription of a relevant drug [15]. A main assumption was that all individuals with T1D were treated with insulin (A10A).

We first conducted analyses restricted to individuals registered in the NDR-A. For these individuals, we extracted all available data from the three administrative registries. We then assessed whether the classification based on the criteria described above corresponded with the diagnosis recorded in the NDR-A. Correspondence was calculated as the overall proportion registered with the same diabetes type in both registries. To evaluate the reliability of diabetes type classification between the NDR-A and the administrative registry data, we employed Cohen’s Kappa statistic. This calculation was performed using the kappa2 function from the irr package in R.

When applying the classification to the whole population aged 18 years or older to estimate prevalence of T1D and T2D in 2022, we included all who fulfilled the criteria based on data registered at any time during 2019-2022 and who were alive and residing in Norway during 2022.

### Human ethics and consent to participate statement

This study is part of the DIANOREG project (Population-based register study of diabetes in Norway). The study was approved by the Regional Committee for Medical and Health Research Ethics (REK 28748 (2019/1175)). Consent to participate was not applicable for this registry-based study.

## Results

Among the 4,315,563 individuals 18 years and older living in Norway during 2022, there were 271,067 individuals who fulfilled the defined criteria for having prevalent diabetes in 2022. Among these, 81,332 individuals were registered in the NDR-A (Figure 1).

### Administrative registry data for individuals registered in the NDR-A

Among those categorized as having T1D in the reference NDR-A, 98.2% had at least one entry of a T1D diagnosis code from specialist care, and 75.9% had at least one entry of a T1D diagnosis from primary care (Table 1).

**Table 1.** Registrations in administrative health registries 2019–2022 for individuals in the Norwegian Diabetes Registry for Adults (NDR-A), by NDR-A diabetes type.

|  | NDR-A diabetes type |  |
| --- | --- | --- |
|  | T1D (n=24,734) | T2D (n=56,121) |
| Registrations in administrative health registries | % | % |
| *NPR (Specialist health care)* |  |  |
| E10 Type 1 diabetes mellitus | 98.2 | 11.1 |
| E11 Type 2 diabetes mellitus | 28.7 | 86.6 |
| *NRPHC (Primary health care)* |  |  |
| T89 Diabetes type 1 | 75.9 | 9.7 |
| T90 Diabetes type 2 | 46.4 | 96.4 |
| *NorPD (Prescribed drugs)* |  |  |
| A10 Drugs use in diabetes | 98.9 | 90.8 |
| A10A Insulins and analogues | 97.6 | 32.9 |
| A10AB Fast-acting insulins | 95.9 | 18.3 |
| A10B Blood glucose lowering drugs, excluding insulins | 12.4 | 87.7 |
| A10BA Metformin | 8.8 | 72.0 |

T1D: Type 1 Diabetes; T2D: Type 2 Diabetes; NPR: The Norwegian Patient Registry; NRPHC: Norwegian Registry for Primary Health Care; NorPD: Norwegian Prescribed Drug Database

Similarly, among those classified as T2D in the reference NDR-A, 86.6% had at least one entry of a T2D diagnosis code from specialist care and 96.4% from primary care.

In those classified as T1D in the reference NDR-A, 97.6% were registered with any insulins or analogues in the NorPD, while 32.9% of those with T2D had used insulin at least once during the study period. In the NDR-A T2D population, 87.7% had used any type of non-insulin blood glucose lowering drugs during the study period.

### Distinguishing T1D and T2D in administrative registry-linkage data

Based on the data in Table 1 and the overall assumption that patients with T1D are treated with insulin, we determined three steps for classification of T1D and T2D based on administrative registry-linkage data.

Figure 2 describes our method for classifying T1D and T2D based on the administrative health registry data for individuals in the NDR-A.

**Figure 2.**
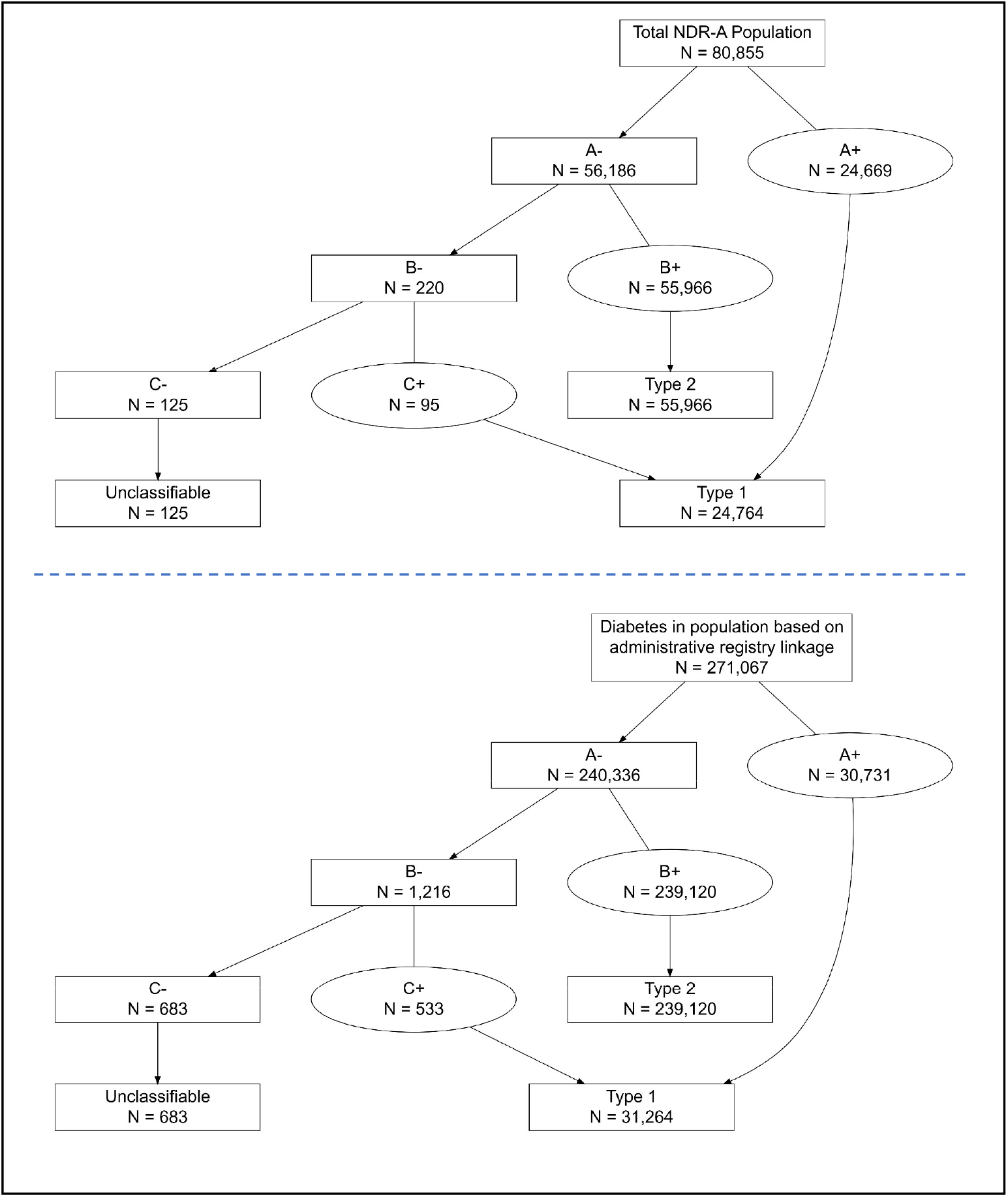
Classification of type 1 (T1D) and type 2 diabetes (T2D) based on administrative registry data. **Top**: Individuals in the reference registry (Norwegian Diabetes Registry for NDR-A). **Bottom**: individuals in the total population with prevalent diabetes, based on administrative registry-linkage data from primary or specialist health care. **Classification step A:** Specialist health care diagnosis for T1D AND insulin AND (never a specialist health care diagnosis for T2D OR never non-insulin glucose lowering drugs OR T1D more than twice as often registered as T2D in specialist health care); **Classification step B**: Diagnosis of T2D in either specialist or primary health care; **Classification step C:** Diagnosis of T1D diagnosis in primary health care AND insulin.

#### Step A, T1D definition

Specialist health care diagnosis T1D (i.e. ICD-10 code E10) *and* insulin treatment (ATC A10A) registered *and*:

- Never a specialist health care diagnosis T2D (i.e. never ICD-10 code E11) (n=17,549) OR
- Never blood glucose lowering drugs other than insulins (i.e. no A10B registrations) (n=5,816) OR
- Specialist health care diagnosis T1D twice or more often registered than specialist health care diagnosis T2D (n=1,304)

Step B, T2D definition: Among the remaining individuals, those with a T2D diagnosis from specialist health care (ICD-10: E11) or from primary health care (ICPC2: T90) were classified as having T2D.

Step C, T1D definition: Among the remaining individuals, those with a T1D diagnosis from primary health care (ICPC2: T89) from *and* insulins were classified as T1D. From the criteria in step B, these individuals never had a T2D diagnosis from either specialist or primary health care.

Following the application of these three classification steps, 125 individuals remained unclassifiable. These individuals had diagnoses of T1D, but no insulin use, and no diagnoses of T2D.

The correspondence for T1D between the classification method and the NDR-A registration was 94.7% and 97.9% for T2D (Table 2). For T1D, the correspondence was lower in the older age-groups, while for T2D, the correspondence was high in all age-groups (Table 2). Overall Cohen’s kappa was 92.7% and was lower in the oldest age groups (18-39 years: 93.4%; 40-49 years: 94.0%, 60-79 years: 87.2% and 80+: 75.0%) with only small differences between the sexes (not shown).

**Table 2.**
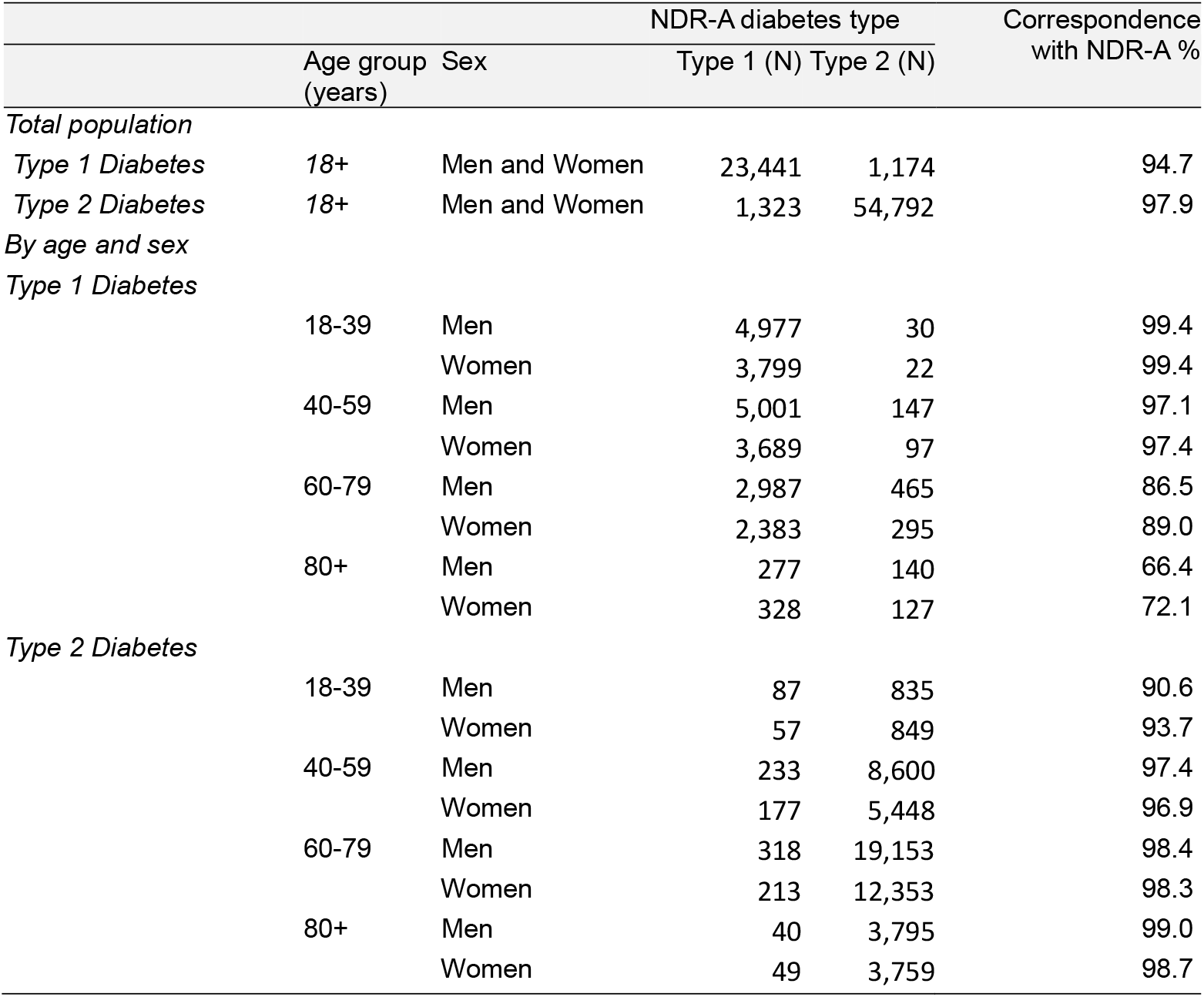
Correspondence between registered diabetes type in the Norwegian Diabetes Registry for Adults (NDR-A) and classification based on administrative health registry data.

### Prevalence of T1D and T2D in Norway in 2022

The final administrative registry-linkage population of 271,067 corresponds to 6.3% of the population 18 years and older in Norway in 2022. By applying the classification to this population, we found that 239,120 individuals (88.2%) were grouped as having T2D, while 31,264 individuals (11.5%) were grouped as having T1D (Figure 2, right panel). Supplemental Table 1 shows the proportion of the population, by sex and age categories, classified as having either T1D or T2D by the administrative registry-linkage.

Table 3 shows the distribution of administrative registry-linkage data for individuals classified as having T1D or T2D (total population). Among those classified as having T1D, 98.3% were registered with a specialist health care diagnosis of T1D, 30.8% were also registered with a specialist health care diagnosis of T2D at least one time. Among those classified as having T1D, 94.0% were registered with fast-acting insulins.

**Table 3.** Registrations in administrative health registries 2019-2022 for individuals in the total population 2022, by diabetes type classification.

| Registrations in administrative health registries | Diabetes type based on<br>administrative health registry data |  |
| --- | --- | --- |
|  | T1D (n=31,264)<br>% | T2D (n=239,120)<br>% |
| *NPR (Specialist health care)* |  |  |
| E10 Type 1 diabetes mellitus | 98.3 | 5.5 |
| E11 Type 2 diabetes mellitus | 30.8 | 69.8 |
| *NRPHC (Primary health care)* |  |  |
| T89 Diabetes type 1 | 72.1 | 5.4 |
| T90 Diabetes type 2 | 48.5 | 94.5 |
| *NorPD (Prescribed drugs)* |  |  |
| A10 Drugs use in diabetes | 100 | 80.0 |
| A10A Insulins and analogues | 100 | 18.2 |
| A10AB Fast-acting insulins | 94.0 | 9.4 |
| A10B Blood glucose lowering drugs, excluding insulins | 14.1 | 78.1 |
| A10BA Metformin | 9.8 | 63.4 |

T1D: Type 1 Diabetes; T2D: Type 2 Diabetes; NPR: The Norwegian Patient Registry; NRPHC: Norwegian Registry for Primary Health Care; NorPD: Norwegian Prescribed Drug Database

The majority of those classified (69.8%) as having T2D were registered with a specialist health care diagnosis of T2D, and 80.0% were dispensed blood glucose lowering drugs (any A10), while 78.1% were treated with non-insulin blood glucose lowering drugs at any point in time during the study period (Table 3). We furthermore observed that 18.2% of those classified as having T2D had used insulin at least one time.

### Robustness analyses

We reanalysed data after reversing the steps, to first identify T2D and then T1D. We found that the correspondence with the classification in the reference NDR-A was similar. We also reanalysed the classification performance with a longer look-back period – the maximum period of available data from each registry and again found similar results (Supplement table S2).

## Discussion

Using the medical quality registry NDR-A as a reference, we found that a classification method combining information from three different nationwide registry sources—covering primary and specialist health care as well as dispensed prescriptions—allowed us to identify diabetes type with high correspondence with the reference registry across most age-groups and for both sexes. By applying this classification method to the total population, we found that in the Norwegian adult population in 2022, 31,264 (0.7%) had T1D while 239,120 (5.5%) had T2D.

### Comparison with other studies

Many different classifications have been used to define diabetes types using administrative data with variable or uncertain validity [4]. Requiring multiple entries of diagnosis codes specific to diabetes type seems to perform well to distinguish between diabetes types in other studies [3, 24, 25]. Previous research has shown that the highest performance is achieved when more than one criterion is applied, for instance diagnosis codes and drugs used in diabetes [4]. Some studies use only blood-glucose lowering drugs to define diabetes cases [26]. In our population, using this approach would have missed 20% of individuals with T2D. In a Norwegian study from a 2014 sample, the proportion of individuals treated with diet only was 31.7% [27], suggesting that the non-pharmacologically treated proportion is declining. Consistently, data from the Swedish National Diabetes Register showed decreasing proportion of non-pharmacologically treated patients from 25% in 2010 to 12% in 2022 [28].

The correspondence between diagnoses from NDR-A and the classification from the linked registry data in our study was comparable to what was reported from Danish registry data by Isaksen et al. for individuals aged 18-74 years in 2018 [14]. Isaksen et al. also combined information on blood-glucose lowering drugs use and diagnosis codes from administrative registries but used self-reported survey data as reference. In their sample of 2633 individuals, the overall estimated positive predictive value (PPV) for T1D was 94%, decreasing to 66% for individuals 40 years or older. The overall PPV for T2D was 90%, dropping to 56% for individuals younger than 40 years.

Prevalence is strongly dependent on the age-groups studied. We published a T2D prevalence of 6.1% in 2014 in those 30 to 89 years in Norway [15]. Similar to our results, a Scottish national report on diabetes in 2022, among all 20 years and older 0.8% had T1D and 7.4% had T2D [29].

### Strengths and limitations

The major strength of this study is the nationwide approach comparing administrative health registry data with data from a large medical quality registry. Our classification method was relatively simple but combined information from diagnosis codes from both primary and secondary care with detailed information on drugs dispensed. Few other countries have nationwide registries for primary care. Using data from primary care is important, as most of the health care contacts for individuals with T2D are in primary care [30].

A limitation of the study is that the same physicians report to NDR-A and to NPR or NRPHC. An important assumption is that completing a form from the NDR-A will be done with greater precision and diligence than entering diagnostic codes for reimbursement purposes. Among the T2D population in the NDR-A, a higher proportion used insulin (33%) compared to those in the administrative registry-linkage population (18%). Additionally, fewer individuals were non-pharmacologically treated in the NDR-A (8.5%) compared to the registry-population (20%). Moreover, a higher proportion with T2D in the NDR-A had been registered in specialist care compared to the total T2D population (90% compared to 75%). This suggests that individuals with T2D in NDR-A are not entirely representative of the total T2D population in Norway. Another limitation is that LADA is grouped together with T1D in the NDR-A, which might explain that 2.4% of individuals classified as T1D (or LADA) in NDR-A did not initiate insulin treatment during the first years after diagnosis.

A problem increasingly facing all use of prescription drugs for classification or identification of diabetes is the use of GLP-1 analogues and SGLT-2 inhibitors for other conditions such as obesity, kidney disease, and heart failure in individuals without diabetes. Metformin can also be prescribed for polycystic ovarian syndrome in some cases. Finally, our estimated prevalence does not account for undiagnosed individuals, who are estimated to represent between 11 - 22% of diabetes cases in Norway.

### Study implications and generalizability

The national administrative health registries collect important data for the entire population and enable the tracking of individuals over time. These registries serve as important sources of information, providing overviews of specific diagnostic groups. However, this benefit is dependent on accurate diagnostic information. Based on our current work, we can use linked data from administrative health registries to conduct epidemiological studies of T1D and T2D. The results show the importance of having access to primary health care data in such studies, especially for T2D. Our results are likely generalizable to settings with access to data from primary health care, specialist health care and prescribed drugs.

### Conclusion

A classification method that combines nationwide information from primary and specialist health care, along with dispensed drugs, was able to correctly identify diabetes type for well above 90% for both T1D and T2D cases.

## Data Availability

The data utilized in this study are derived from Norwegian health registries and contain sensitive personal information. Access to these data is available to researchers through an application process described at www.helsedata.no.

## Authorship contribution statement

The study was conceptualized by PLD, IJB, LCS and HLG, and administered by HLG and KFL. Figure 1 was prepared by PLR. IJB handled and analysed the data and prepared figures 2 and 3. The first draft of the main manuscript text was written by PLD, LCS and IJB. All authors provided substantial contributions to the interpretation of data, reviewed the manuscript critically, approved the final version to be published and agreed to be accountable for all aspects of the manuscript. All authors reviewed and edited the manuscript and approved the submitted version.

## Disclaimer

Data from the Norwegian Patient Registry and the Norwegian Registry of Primary Health Care have been used in this publication. The interpretation and reporting of these data are the sole responsibility of the authors, and no endorsement by the Norwegian Patient Registry or the Norwegian Registry of Primary Health Care is intended or should be inferred.

