## Supplementary Material for "Classification of type 1 diabetes and type 2 diabetes based on administrative registry data: A nationwide study from Norway"

"Validation of algorithms for diabetes classification based on linked register-data in Norway"

**Supplemental Table S1. Prevalence of diabetes in Norway 2022, stratified by age and sex.**

| | N (Population Percentage) | |
|---|---|---|
| | T1D | T2D |
| **Total** | | |
| | 31,264 (0.7%) | 239,120 (5.5%) |
| **All ages** | | |
| Men | 17,944 (0.8%) | 138,534 (6.4%) |
| Women | 13,320 (0.6%) | 100,586 (4.7%) |
| **18-39** | | |
| Men | 5,701 (0.7%) | 3,809 (0.5%) |
| Women | 4,251 (0.6%) | 3,551 (0.5%) |
| **40-59** | | |
| Men | 6,339 (0.9%) | 36,928 (5.1%) |
| Women | 4,450 (0.6%) | 24,196 (3.5%) |
| **60-79** | | |
| Men | 5,040 (0.9%) | 78,956 (14.7%) |
| Women | 3,730 (0.7%) | 52,961 (9.7%) |
| **80+** | | |
| Men | 864 (0.8%) | 18,841 (16.9%) |
| Women | 889 (0.6%) | 19,878 (12.4%) |

**Supplemental Table S2. Algorithm performance (sensitivity and PPV) from administrative register-linkage data to NDR-A using all available look-back period in the registers** (NDR-A from 2009, NPR from 2008, KPR from July 2016 and LMR from 2004).

| | | NDR-A (reference population) | | Algorithm's performance | |
|---|---|---|---|---|---|
| | | T1D | T2D | Sensitivity (%) | PPV (%) |
| **Type 1 Diabetes** | | | | | |
| **18-39** | | | | | |
| | M | 5,130 | 32 | 98.5 | 99.4 |
| | F | 3,881 | 24 | 98.4 | 99.4 |
| **40-59** | | | | | |
| | M | 5,124 | 128 | 93.6 | 97.6 |
| | F | 3,789 | 82 | 94.1 | 97.9 |
| **60-79** | | | | | |
| | M | 2,958 | 366 | 84.8 | 89.0 |
| | F | 2,374 | 225 | 86.6 | 91.3 |
| **80+** | | | | | |
| | M | 276 | 143 | 77.3 | 65.9 |
| | F | 317 | 141 | 76.0 | 69.2 |
| **Type 2 Diabetes** | | | | | |
| **18-39** | | | | | |
| | M | 79 | 996 | 96.9 | 92.7 |
| | F | 63 | 976 | 97.6 | 93.9 |
| **40-59** | | | | | |
| | M | 352 | 10,416 | 98.8 | 96.7 |
| | F | 239 | 6,655 | 98.8 | 96.5 |
| **60-79** | | | | | |
| | M | 531 | 24,147 | 98.5 | 97.8 |
| | F | 367 | 15,723 | 98.6 | 97.7 |
| **80+** | | | | | |
| | M | 81 | 5,302 | 97.4 | 98.5 |
| | F | 100 | 5,357 | 97.4 | 98.2 |

*Note:*
A total of 107 persons not classified by the algorithm are excluded
